# Derivation and validation of a machine learning risk score using biomarker and electronic patient data to predict rapid progression of diabetic kidney disease

**DOI:** 10.1101/2020.06.01.20119552

**Authors:** Lili Chan, Girish N. Nadkarni, Fergus Fleming, James R. McCullough, Patti Connolly, Gohar Mosoyan, Fadi El Salem, Michael W. Kattan, Joseph A. Vassalotti, Barbara Murphy, Michael J. Donovan, Steven G. Coca, Scott Damrauer

## Abstract

**Importance:** Diabetic kidney disease (DKD) is the leading cause of kidney failure in the United States and predicting progression is necessary for improving outcomes.

**Objective:** To develop and validate a machine-learned, prognostic risk score (KidneyIntelX™) combining data from electronic health records (EHR) and circulating biomarkers to predict DKD progression.

**Design:** Observational cohort study

**Setting:** Two EHR linked biobanks: Mount Sinai BioMe Biobank and the Penn Medicine Biobank.

**Participants:** Patients with prevalent DKD (G3a-G3b with all grades of albuminuria (A1-A3) and G1 & G2 with A2-A3 level albuminuria) and banked plasma.

**Main outcomes and measures:** Plasma biomarkers soluble tumor necrosis factor 1/2 (sTNFR1, sTNFR2) and kidney injury molecule-1 (KIM-1) were measured at baseline. Patients were divided into derivation [60%] and validation sets [40%]. The composite primary end point, progressive decline in kidney function, including the following: rapid kidney function decline (RKFD) (estimated glomerular filtration rate (eGFR) decline of ≥5 ml/min/1.73m^2^/year), ≥40% sustained decline, or kidney failure within 5 years. A machine learning model (random forest) was trained and performance assessed using standard metrics.

**Results:** In 1146 patients with DKD the median age was 63, 51% were female, median baseline eGFR was 54 ml/min/1.73 m^2^, urine albumin to creatinine ratio (uACR) was 61 mg/g, and follow-up was 4.3 years. 241 patients (21%) experienced progressive decline in kidney function. On 10-fold cross validation in the derivation set (n=686), the risk model had an area under the curve (AUC) of 0.77 (95% CI 0.74-0.79). In validation (n=460), the AUC was 0.77 (95% CI 0.76-0.79). By comparison, the AUC for an optimized clinical model was 0.62 (95% CI 0.61-0.63) in derivation and 0.61 (95% CI 0.60-0.63) in validation. Using cutoffs from derivation, KidneyIntelX stratified 46%, 37% and 16.5% of validation cohort into low-, intermediate- and high-risk groups, with a positive predictive value (PPV) of 62% (vs. PPV of 37% for the clinical model and 40% for KDIGO; p < 0.001) in the high-risk group and a negative predictive value (NPV) of 91% in the low-risk group. The net reclassification index for events into high-risk group was 41% (p<0.05).

**Conclusions and Relevance:** A machine learned model combining plasma biomarkers and EHR data improved prediction of progressive decline in kidney function within 5 years over KDIGO and standard clinical models in patients with early DKD.

## INTRODUCTION

Approximately 1 out of 4 adults with type 2 diabetes mellitus (T2D) has kidney disease (i.e. Diabetic Kidney Disease or DKD). Each year, 50,000 individuals with DKD progress to kidney failure in the United States.^1^ The Mount Sinai Health System alone provides care for over 70,000 patients with DKD. Estimated glomerular filtration rate (eGFR) and urinary albumin creatinine ratio (uACR), existing diagnostic measurements incorporated into the Kidney Disease: Improving Global Outcomes (KDIGO) guidelines for risk stratification,^2^ lack precision in identifying patients who will experience rapid kidney function decline (RKFD), especially in earlier stages of DKD (G1-G3).^3^ As a result, primary care physicians (PCP) and diabetologists are often not able to appropriately risk stratify and counsel patients on the progressive nature of their disease. Easily interpretable and accurate prognostic tools that integrate into clinical workflow are lacking, resulting in suboptimal treatment and referral delays to a nephrology specialist. This has led, in part, to the unacceptable level of RKFD (and kidney failure) in this population^4-8^ with a high proportion of patients starting unplanned dialysis.^1,9,10^

Several blood-based biomarkers have shown associations with DKD progression, most significantly soluble tumor necrosis factor receptors 1/2 (TNFR1/2), and plasma kidney injury molecule-1 (KIM-1).^11-13^ However, implementation of accurate prognostic models combining clinical data from patients’ electronic health record (EHR) with blood-based biomarkers is lacking. Although EHR data is widely available, its volume and complexity limits integration with biomarker values using traditional methodologies. Recently, machine learning approaches have been developed that can combine biomarkers and EHR data to produce prognostic risk scores. A simple risk score that improves the ability to identify patients with DKD at low, intermediate, and high risk of RKFD has the potential to improve outcomes through more effective use of medications and efficient resource allocation at the primary care physician level.

In the current study, we developed and validated the performance of a biomarker-enriched, machine learned risk score (i.e., the KidneyIntelX™ test) to predict progressive decline in kidney function in patients with early stage DKD and compared performance to standard clinical models. We determined risk-based thresholds that can easily be integrated into standard clinical workflows and enhance existing clinical practice guidelines.

## MATERIALS AND METHODS

### Study Sample

The cohort is derived from the BioMe Biobank at the Icahn School of Medicine at Mount Sinai (ISMMS) and Penn Medicine Biobank (PMBB). The BioMe Biobank is a plasma and DNA biorepository with recruitment from 2007 which includes informed-consent access to the patients’ EHR from a diverse local community in New York City.^14,15^ PMBB is a research cohort enrolled from the University of Pennsylvania Health System with recruitment from 2008.^14^ Participants actively consented to allow the linkage of biospecimens with their longitudinal EHR (**eFigure 1**). Both BioMe and PMBB are institutional biobanks that attempt to be representative of the patient populations of the institutions they serve. Patients are recruited from outpatient general medicine clinics and certain subspecialty clinics with limited pre-selection criteria.^16,17^

The study protocol was approved by institutional review boards at both ISMMS and University of Pennsylvania; all participants had provided broad written informed consent for research and were not specifically compensated for participation in the current study. Blood was collected on the day of enrollment into BioMe or PMBB with plasma isolation as per standard procedures and continuously stored at −80°C until shipping to the RenalytixAI laboratory, New York, NY where biomarker measurements were performed.

### Inclusion Criteria

We selected patients from BioMe and PMBB who were 21-81 years at the time of biobank enrollment (“baseline”), with T2D, an eGFR between 30 and 59.9 ml/min/1.73m^2^ or an eGFR≥60 ml/min/1.73 m^2^ with uACR ≥ 30mg/g. The KDIGO risk model categorizes patients based on eGFR and albuminuria and has 3 colors that correspond to the prognosis of prevalent CKD (we did not include patients at “low risk” or green because they do not have CKD).^2^ Patients were included if by the KDIGO eGFR and uACR criteria they were stage G3a-G3b with all grades of albuminuria (A1-A3) and G1, G2 with moderate to high albuminuria (UACR ≥ 30 mg/g (A2-A3)).^2^ Proportion of each DKD stage was evaluated against national estimates derived from the National Health and Nutrition Examination Survey (NHANES) years 2018-2019.^18^ For eGFR, we defined the baseline period as 1 year before or up to 3 months after the biobank enrolment date. Baseline uACR values were derived from closest values ±1 year from enrolment to maximize sample size as these are measured less frequently; subjects without baseline values of eGFR and uACR meeting these criteria were excluded. Only patients with a stored plasma specimen, a minimum follow-up time from enrolment of at least 21 months, and ≥3 eGFR values after baseline (**eFigure 1**) were included. Patients with kidney transplants or on chronic maintenance dialysis before baseline were excluded from the study.

### Ascertainment of clinical variables

#### Bio*Me* Biobank and PMBB

Sex and race were obtained from biobank questionnaires or EHR data. Clinical data was extracted for all EHR variables with concordant time stamps. Hypertension and T2D status at baseline were determined using the eMERGE Network phenotyping algorithms.^16^ Cardiovascular disease and heart failure were determined by International classification of disease (ICD)-9/10 clinical modification codes.

#### Biomarker Assays

The three plasma biomarkers were measured in a proprietary, analytically validated multiplex format using the Mesoscale platform (MesoScale Diagnostics, Gaithersburg, Maryland, USA), which employs electrochemiluminescence detection methods combined with patterned arrays to allow for multiplexing of assays. Each sample was run in duplicate, along with quality control samples with known low, moderate, and high concentrations of each biomarker on each plate. Assay precision was assessed using a panel of 7 reference samples that spanned the measurement range. The intra-assay coefficient of variation (CV) results for KIM-1, TNFR1, and TNFR2 were mean CV 3.9%, 5.4%, and 3.7%, respectively. The inter-assay CV results for the reference samples for KIM-1, TNFR-I, and TNFR-2 were mean CV 9.9%, 10.1%, and 7.8%, respectively. Assays satisfied dilution linearity and were run at 1:4 dilution. Levey-Jennings plots were employed and followed the Westguard rules for re-run of samples. The laboratory personnel performing the biomarker assays were blinded to all clinical information.

#### Data Harmonization

We harmonized data from Bio*M*e and PMBB. Race/ethnicity was collapsed into 4 major, non-overlapping categories (White, Non-Hispanic Black, Hispanic, and Other). ICD and Current Procedural Terminology (CPT) codes were included as yes/no variables with timestamps. Medications were mapped to RxNorm codes^19^ and laboratory values to Logical Observation Identifiers Names and Codes (LOINC) codes.^20^ Only variables represented in >70% of subjects throughout the combined dataset (except uACR and blood pressure due to their established clinical importance) were included and used for training of the KidneyIntelX algorithm.

#### Ascertainment and definition of the kidney endpoint

We determined eGFR using the CKD-EPI creatinine equation.^21^ We employed linear mixed models with an unstructured variance-covariance matrix and random intercept/slope for each individual to estimate eGFR slope.^22^ The primary composite outcome, progressive decline in kidney function, included the following: RKFD defined as an eGFR slope decline of ≥ 5 ml/min/1.73 m^2^/year,^2^ a sustained (confirmed at least 3 months later) decline in eGFR of ≥40%^23^ from baseline, or “kidney failure” defined by sustained eGFR < 15 ml/min/1.73 m^2^ confirmed at least 30 days later, or receipt of long-term maintenance dialysis or receipt of a kidney transplant.^2^ Additionally, two nephrologists (SC/GNN) independently adjudicated all outcomes examining each individual patient over their longitudinal course, accounting for eGFR changes (ensuring annualized decline of ≥5 ml/min or ≥ 40% sustained decrease), corresponding ICD/CPT codes and medications to ensure that outcomes represented true decline rather than a context dependent temporary change (e.g., due to medications/hospitalizations). Follow up time was censored after loss to follow-up, after the date that the non-slope components of the composite kidney endpoint were met, or 5 years after baseline.

#### Statistical Analysis

The datasets were randomized into a derivation (60%) and validation sets (40%). The validation dataset was completely blinded and sequestered from the total derivation dataset. Using only the derivation set, we evaluated supervised random forest algorithms on the combined biomarker and all structured EHR features without a priori feature selection and identified a candidate feature set; eTable 1. The derivation set was then randomly split into secondary training and test sets for model optimization with 70%-30% spitting and a 10-fold cross-validation for AUC. We considered both raw values and ratios of the biomarkers. Missing uACR values were imputed to 10 mg/g,^24^ missing blood pressure (BP) values were imputed using multiple predictors (age, sex, race and antihypertensive medications),^25^ and median value was used for other features where missingness was < 30% (**eTable 2**). We conducted further iterations of the model by tuning the individual hyperparameters. A hyperparameter is a parameter which is used to control the learning process (e.g., no of RF trees) as opposed to parameters whose weights are learned during the training (e.g.: weight of a variable). Tuning hyperparameters refers to iteration of model architecture after setting parameter weights to achieve the ideal performance. For random forest architecture, it could include components such as maximum depth of decision tree, number of trees in forest, and majority voting rules.^26^ The final model was selected based on AUC performance.

We generated risk probabilities for the composite kidney endpoint using the final model in the derivation set, scaled them to align with a continuous score from 5-100 by increments of 5, and applied this score to the validation set. Risk cut-offs were chosen in the derivation set to encompass the top 15% as the high risk (scores 90-100), bottom 45% as the low risk (scores 5-45), and the intervening 40% as the intermediate risk group (scores 50-85). Primary performance criteria were AUC, positive predictive value for high risk group and negative predictive values for low risk group (PPV and NPV, respectively) at the pre-determined cut-offs. The selected model and associated cut-offs were then validated by an independent biostatistician (MK) in the sequestered validation cohort.

In addition to these traditional test statistics, we assessed calibration by examination of the slope of observed vs. expected outcome plots of the KidneyIntelX score vs. only the observed outcomes. We also constructed Kaplan Meier curves for time-dependent outcomes of 40% decline and kidney failure with hazard ratios using the Cox proportional hazards method.

The discrimination of the KidneyIntelX model was compared to a recently validated comprehensive clinical model which included age, sex, race, eGFR, cardiovascular disease, smoking, hypertension, BMI, UACR, insulin, diabetes medications, and HbA1c and was developed to predict 40% eGFR decline in eGFR in T2D.^24^ Utility metrics (PPV, NPV) were compared to both the comprehensive clinical model and KDIGO risk strata. Finally, we calculated the net reclassification index (NRI) for events and non-events compared to KDIGO risk strata.^27,28^ All a-priori levels of significance were <0.05. All hypothesis tests were two-sided. All analyses were performed with R software (www.rproject.org).

## RESULTS

### Baseline Characteristics of Cohorts

Baseline characteristics of the total study cohort incorporating derivation and validation (n=1146) were as follows; median age 63 years, 581 (51%) female, median eGFR was 54 ml/min/1.73 m^2^, and the median uACR was 61 mg/g. uACR was available in 62% of the cohort and imputed to 10 mg/g in 38%. The most common comorbidities were hypertension (91%), coronary heart disease (35%), and heart failure (33%). The majority (81%) were on ACE inhibitors or angiotensin receptor blockers (ARBs). Baseline characteristics between derivation and validation sets including event rates were balanced. The median number of serum creatinine/eGFR values per patient during the follow-up period was 16 (**Table 1**). Distribution of DKD stages of the study cohort is similar to national estimates (**eTable 3**).

**Table 1.** Clinical Characteristics of the Participants in the Derivation and Validation Cohorts.

|  | Study Population<br>n = 1146 | Derivation Population<br>n = 686 | Validation Population<br>n = 460 |
| --- | --- | --- | --- |
| **Clinical Characteristic - Median [Q1 - Q3]** |  |  |  |
| Age in years, Median [IQR] | 63 [55 - 69] | 63 [55 - 68] | 63 [56 - 69] |
| Female, n (%) | 581 (50.7) | 352 (51.3) | 229 (49.8) |
| **Race, n (%)** |  |  |  |
| White | 373 (32.6) | 231 (33.7) | 142 (30.8) |
| African-American | 386 (33.7) | 226 (32.9) | 160 (34.8) |
| Others | 387 (33.8) | 229 (33.4) | 158 (34) |
| Body Mass Index, Median [IQR] | 31 [29 - 35] | 31 [29 - 35] | 31 [29 - 36] |
| Hypertension, n (%) | 1043 (91.0) | 622 (90.7) | 421 (91.5) |
| Coronary Artery Disease, n (%) | 406 (35.4) | 234 (34.1) | 172 (37.4) |
| Heart Failure, n (%) | 378 (33) | 213 (31.1) | 165 (35.9) |
| Systolic BP in mm of Hg, Median [IQR] | 130 [120 - 144] | 130 [119 - 144] | 130 [120 - 144] |
| Diastolic BP in mm of Hg, Median [IQR] | 74 [67 - 81] | 74 [66 - 81] | 73 [67 - 80] |
| Mean Arterial Pressure in mm of Hg, Median [IQR] | 93.3 [85.7 - 100.7] | 93.3 [85.1 - 100.7] | 93.3 [86 - 100.4] |
| Follow Up Time In Months, Median [IQR] | 51.9 [36.5 - 58.1] | 51.3 [36.8 - 58.1] | 52.8 [35.9 - 58.1] |
| **Laboratory Characteristics - Median [Q1 - Q3]** |  |  |  |
| Baseline estimated glomerular filtration rate in ml/min, Median [IQR] | 54.3 [45.3 - 67.3] | 54.4 [44.4 - 68.4] | 54.1 [45.7 - 66.1] |
| **eGFR Strata** |  |  |  |
| [30 - 44.9] (n, %) | 279 (24.4) | 176 (25.7) | 103 (22.4) |
| [45 - 59.9] (n, %) | 490 (42.8) | 275 (40.1) | 215 (46.7) |
| [60 - 89.9] (n, %) | 263 (22.9) | 170 (24.8) | 93 (20.2) |
| [>=90] (n, %) | 114 (9.9) | 65 (9.5) | 49 (10.7) |
| Baseline urine albumin creatinine ratio, Median [IQR] | 61.3 [16 - 241] | 65 [18 - 238] | 53.5 [15 - 246.3] |
| Missing urine albumin creatinine ratio, n (%) | 433 (37.8) | 269 (39.2) | 164 (35.6) |
| Baseline Hemoglobin A1C in %, Median [IQR] | 6.9 [6.2 - 8.2] | 7 [6.2 - 8.22] | 6.9 [6.2 - 8.2] |
| **Medication** |  |  |  |
| Angiotensin Converting Enzyme/Angiotensinogen Receptor Blocker, n (%) | 926 (80.8) | 560 (81.6) | 366 (79.6) |
| **Biomarkers pg/ml- Median [Q1 - Q3]** |  |  |  |
| TNFR1 | 2807 [2192 - 3830] | 2807 [2191 - 3830] | 2924 [2217-3894] |
| TNFR2 | 11090 [8031 - 14984] | 11090 [8031 - 14984] | 11171 [8302-15046] |
| KIM-1 | 124 [76 - 235] | 124 [76 - 235] | 138 [82-253] |
| **Smoking Status** |  |  |  |
| Never, n (%) | 354 (30.9) | 214 (31.2) | 140 (30.4) |
| Ever, n (%) | 503 (43.9) | 298 (43.4) | 205 (44.6) |
| Missing, n (%) | 289 (25.2) | 174 (25.4) | 115 (25) |
| **Events\*** |  |  |  |
| eGFR slope $\geq 5$ ml/min/1.73 m<sup>2</sup>/year, n (%) | 171 (14.9) | 98 (14.2) | 73 (15.9) |
| Sustained 40% Decline in eGFR, n (%)<sup>a</sup> | 179 (15.6) | 103 (15) | 76 (16.5) |
| Kidney Failure, n (%)<sup>b</sup> | 52 (4.5) | 29 (4.2) | 23 (5) |
| Composite Endpoint, n (%)<sup>c</sup> | 241 (21) | 137 (20) | 104 (22.6) |

Definitions:
<sup>a</sup> Sustained 40% decline in eGFR (confirmed at least 3 months later) decline in eGFR of $\geq 40\%$ from baseline
<sup>b</sup> "Kidney failure" defined by sustained eGFR $< 15$ confirmed at least 30 days later, or receipt of long-term maintenance dialysis or receipt of a kidney transplant
<sup>c</sup> Composite Event: Progressive decline in kidney function defined by any of the following: eGFR slope $\geq 5$ ml/min/1.73 m<sup>2</sup>/year or sustained 40% decline in eGFR or kidney failure
Abbreviations: TNFR1-tumor necrosis factor 1; TNFR2- tumor necrosis factor 2; KIM-1- kidney injury molecule-1

### Prediction of the composite kidney endpoint (progressive decline in kidney function)

Overall, 241 patients (21%) experienced progressive decline in kidney function over a median 4.3 (IQR 3.0-4.8) years. In the complete derivation set (n=686), using 10-fold cross validation for discrimination, the average AUC for the KidneyIntelX model was 0.77 (95% CI 0.74-0.79). The most significant data features contributing to performance of the KidneyIntelX model included the three plasma biomarkers (TNFR1, TNFR2 and KIM1, as discrete values and ratios), eGFR, uACR, and systolic blood pressure (**Figure 1**). This final model had an AUC of 0.77 (95% CI 0.76-0.79) in the validation set (n=460). The risk for the composite kidney event increased by predicted probabilities of the KidneyIntelX score (**Figures 2a** and **eTable 4**) and by the KidneyIntelX score (**Figure 2b** and **eTable 4**). The slope of the observed vs. the predicted risk for KidneyIntelX was 0.8 in the training set and 1.0 in the validation set, indicating good calibration (**eFigure 2**). By comparison, the comprehensive clinical model yielded an AUC of 0.62 (95% CI 0.61-0.63) in the full derivation set (n=686) and 0.61 (95% CI 0.60-0.63) in validation set (n=460); Delong p value for KidneyIntelX vs. clinical model <0.001).

**Figure 1.**
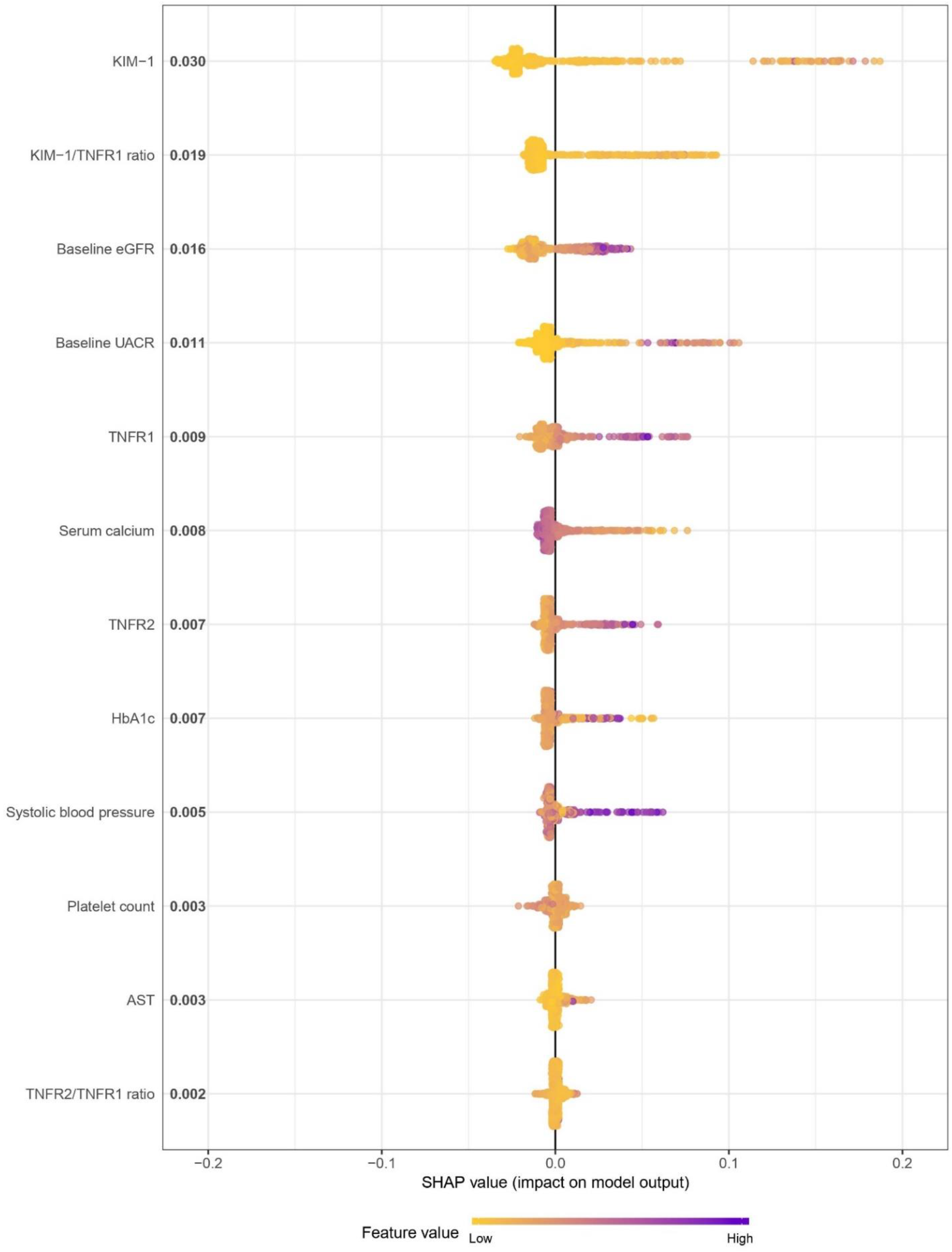
Shapley additive explanations (SHAP) plot showing relative feature importance

**Figure 2.**
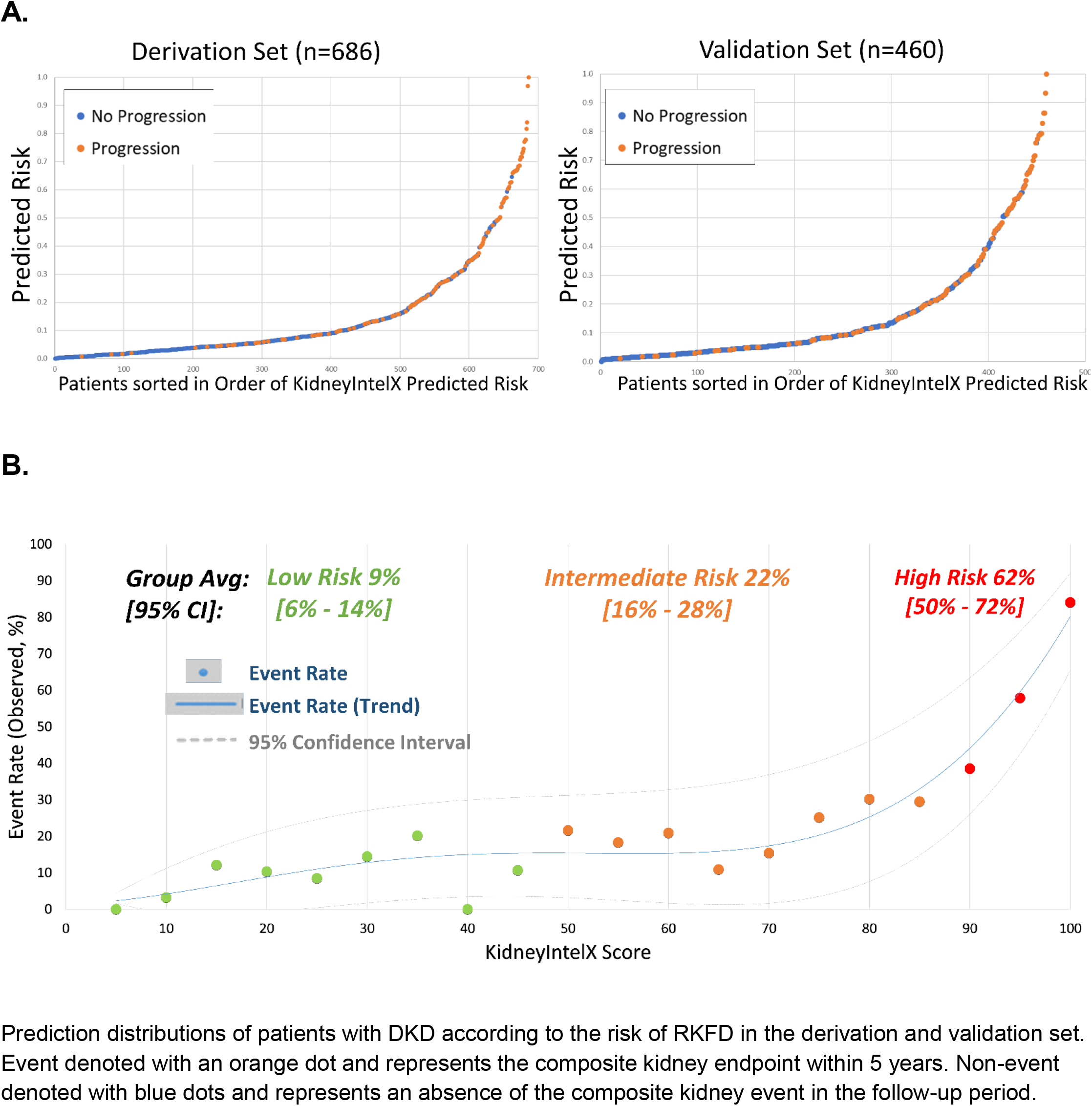
Composite Kidney Endpoint Event Rates by A. KidneyIntelX Predicted Risk and B. KidneyIntelX Score

### KidneyIntelX Clinical Utility Cut-points

The risk probability cutoffs of KidneyIntelX selected in the derivation set (n=686) were 0.061 for the lowest 45% of patients and 0.302 for the top 15% of patients. When these risk cut-offs were applied to the complete validation set, with imputed uACR for missing values (n=460), KidneyIntelX stratified patients to low- (46%), intermediate- (37%), and high-risk (16.5%) groups with respective probabilities for the composite kidney endpoint of 0.09, 0.22, and 0.62. When the optimized clinical model was applied to the validation set, the respective probabilities for the composite kidney endpoint was 0.171 for the bottom 46% of the population and 0. 319 for the top 16.5%. Thus, the PPV was 62% in the KidneyIntelX high-risk group compared to a PPV of 37% for the comprehensive clinical model, p value < 0.001; Table 2). The NPV in the KidneyIntelX low-risk group was 91% compared to an NPV of 88% for the comprehensive clinical model (p= 0.33). The distribution of patients into KDIGO risk categories was established using 296 subjects (64%) with uACR available in the validation cohort and stratified the population into “moderately increased risk” (53%), “high risk” (31%), and “very high risk” (16%) with respective probabilities of 0.15, 0.29 and 0.40 for the composite kidney endpoint over 5 years. In the subgroup with non-imputed uACR (n=296), the PPV for the high-risk strata of KidneyIntelX was 69% and the NPV for the low-risk strata of KidneyIntelX was 93%. Confusion matrices are available in eTable 5. Additional risk cutoffs by potentially relevant proportions of the population are shown in Table 2 and comparisons to the KDIGO risk strata are in **eTable 6**.

**Table 2.** Test Characteristics for KidneyIntelX and the Comprehensive Clinical Model.

| Predicted Risk | KidneyIntelX Risk Score | Full Derivation Set (n=686)<sup>a</sup> |  |  |  | Validation Set (n=460)<sup>b</sup> |  |  |  | Predicted risk | Optimized Clinical Model |  |  |  |
| --- | --- | --- | --- | --- | --- | --- | --- | --- | --- | --- | --- | --- | --- | --- |
| Low Risk |  | Population | Sens | Spec | NPV | Population | Sens | Spec | NPV | Low Risk | Population | Sens | Spec | NPV |
| **0.040** | ≤30 | Lowest 30% | 96% | 37% | 98% | Lowest 32% | 88% | 38% | 91% | **0.142** | Lowest 32% | 74% | 33% | 86% |
| **0.061** | ≤45 | Lowest 45% | 88% | 53% | 95% | Lowest 46% | 81% | 54% | 91% | **0.171** | Lowest 46% | 67% | 48% | 88% |
| **0.0712** | ≤50 | Lowest 50% | 85% | 59% | 94% | Lowest 48% | 77% | 58% | 90% | **0.175** | Lowest 48% | 67% | 51% | 89% |
| High Risk |  | Population | Sens | Spec | PPV | Population | Sens | Spec | PPV | High Risk | Population | Sens | Spec | PPV |
| **0.241** | ≥85 | Top 20% | 56% | 89% | 56% | Top 21% | 50% | 88% | 55% | **0.288** | Top 21% | 41% | 82% | 31% |
| **0.302** | ≥90 | Top 15% | 46% | 93% | 63% | Top 16.5% | 45% | 93% | 62% | **0.319** | Top 16.5% | 37% | 88% | 37% |
| **0.401** | ≥95 | Top 10% | 32% | 96% | 67% | Top 12% | 31% | 96% | 70% | **0.361** | Top 12% | 28% | 91% | 38% |

Abbreviations: Sens= sensitivity, spec= specificity, PPV= positive predictive value; NPV= negative predictive value
<sup>a</sup>AUCs in Derivation Set: 0.85 (95% CI 0.84-0.86) in train and AUC 0.77 (95% CI 0.74-0.79) from 10-fold cross validation testing
<sup>b</sup>AUC in validation set 0.77 (95% CI 0.76-0.79)

KidneyIntelX scores correctly classified more cases into the appropriate risk strata (NRI_event_ = 55% in the derivation set and 41% in the validation set, p value < 0.05; **eTable 7**) compared to KDIGO risk strata. NRI_non-event_ was −8.2% in the derivation set and −7.9% in the validation set (p value NS).

### Supplementary Analyses

#### Time to Event Analyses for 40% Sustained Decline or Kidney Failure

Patients with high-risk KidneyIntelX scores (top 15% in the derivation set and top 16.5% in the validation set) had greater risk of progression to time-to-event categorical outcomes of 40% sustained decline or kidney failure than patients in the low- or medium-risk strata combined (hazard ratio (HR) 9.2; 95% CI: 6.2-13.6 in derivation and 9.1, 95% CI 5.8-14.4 in the validation set; **Figure 3A & 3B**). Kaplan-Meier curves by KDIGO risk categories in the training and validation set are shown in **eFigure 3**.

**Figure 3.**
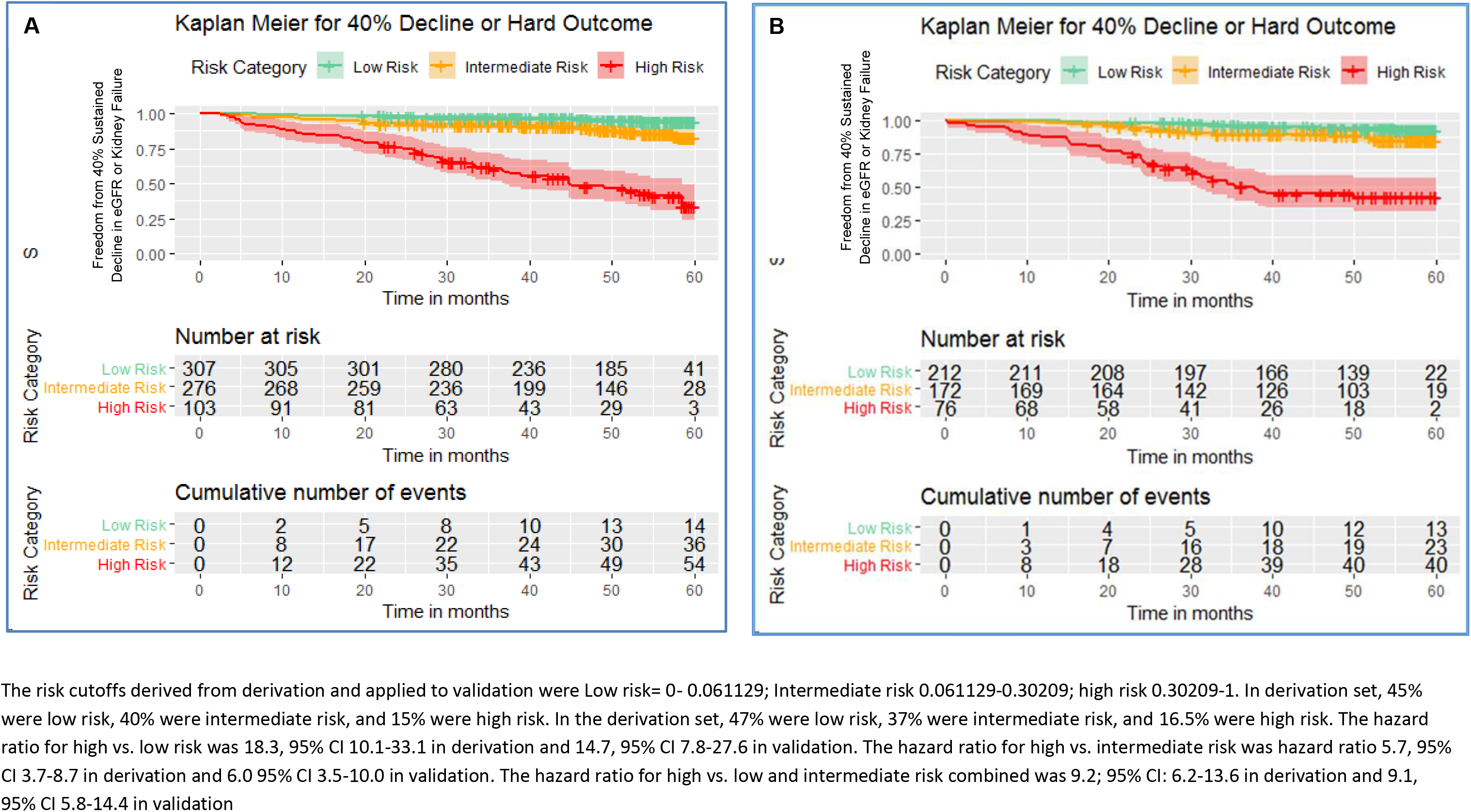
Kaplan-Meier Curves by KidneyIntelX Risk Strata for the Endpoint of Sustained 40% Decline in eGFR or Kidney Failure in Derivation (Panel A) and Validation (Panel B) Sets

#### Subgroup analysis

KidneyIntelX performed similarly across patients with an eGFR greater or less than 60 ml/min/1.73m^2^ at baseline (0.78 and 0.76 respectively). Additionally, when only data in the year prior to enrollment was included, the AUC was identical (0.77) as was the PPV for the top 16% (62%) and the NPV for the bottom 45% (91%). Kaplan-Meier plots did not change when limited to patients with data ≥5 years to ensure alive for at least 5 years (**eFigure 4**).

#### Discrimination for “Kidney Failure” Endpoint

Using the same KidneyIntelX model specifically trained for the composite kidney endpoint, the AUC of KidneyIntelX risk scores for the “kidney failure” endpoint alone was 0.87 (95% CI 0.84-0.89) in the derivation cohort and 0.89 (95% CI 0.87-0.91) in the validation cohort.

## DISCUSSION

Utilizing patients with T2D from two biobanks with plasma samples and linked EHR data, we developed and validated a risk score combining clinical data and three plasma biomarkers via a random forest algorithm to predict a composite kidney outcome, progressive decline in kidney function, consisting of RKFD, sustained 40% decline in eGFR, and kidney failure over 5 years. We demonstrated that the KidneyIntelX outperformed models using only standard clinical variables, including KDIGO risk categories.^3,20^ There were marked improvements in discrimination over clinical models, as measured by AUC, NRI, and improvements in PPV compared to KDIGO risk categories. Furthermore, we showed that KidneyIntelX accurately identified over 40% more patients experiencing events than the KDIGO strata. Finally, KidneyIntelX provided good risk-stratification for the accepted FDA endpoint of sustained 40% decline in eGFR or kidney failure with a 9-fold difference in risk between the high-risk and low- and intermediate-risk strata for this clinical and objective endpoint.

DKD is an increasingly complex and common problem challenging modern healthcare systems. In real world practice, the prediction of RKFD in patients with T2D is challenging, particularly in early disease with preserved kidney function and therefore, implementation of improved prognostic tests is paramount. Our integrated risk score has near-term clinical implications, especially when linked to clinical decision support (CDS) and embedded care pathways. The current standard for clinical risk stratification (KDIGO risk strata)^2^ has three risk strata that overlap with the population of DKD patients that we included in our study. We also created a risk score with three risk strata (low, intermediate, and high) incorporating KDIGO classification components (eGFR and uACR), as well as the addition of other clinical variables, and three blood-based biomarkers. In this way, we were able to augment the ability to accurately risk-stratify DKD patients, thereby enabling improved patient management.

Low- and intermediate-risk patients with DKD can continue care with their existing PCP’s or diabetologists and require less intensity of treatments, unless repeat testing, changes in clinical status or local arrangements regarding referral to specialist care indicate otherwise. For those with high-risk scores, oversight may include more referrals to nephrology,^29,30^ increased monitoring intervals, improved awareness of kidney health, referral to dieticians, reinforcement of usage of antagonists of the renin angiotensin aldosterone system, and increased motivation to start recently approved medications, including SGLT2 inhibitors and GLP-1 receptor agonists to slow progression.^31-34^ Adoption of these new therapies is lagging, especially in patients considered to be ‘low-risk’ by standard criteria, where cost of treatment and presence of adverse events are limiting factors. Earlier engagement with nephrologists may also allow for more time to advise and educate patients about home-based dialysis and pre-emptive or early kidney transplant as patient-centered kidney replacement options if more aggressive treatment does not ultimately prevent progression of DKD. The use of a risk score as part of the enrollment process in future RCTs may enrich the trial participants for greater likelihood of events and thus reduce the chances for type 2 error, or minimize the sample size needed to detect a statistically significant difference with treatment vs. control. Interventions that prevent or slow CKD progression and foster patient-centered kidney replacement modalities support the goals of the US Department of Health and Human Services’ Advancing American Kidney Health initiative.^35^

KidneyIntelX included inputs from biomarkers examined in several settings, including patients with DKD. Soluble TNFR1 and 2 and plasma KIM-1 have demonstrated reliable independent prognostic signals for kidney function decline and ESRD.^11,12,15,36-41^ By incorporating biomarker levels and the EHR data into our machine learning algorithm, we were able to provide a multidimensional representation of the patient and allow for the model to generate improved prognostic estimates.^42,43^

Our study should be interpreted in light of the following limitations. uACR was missing in 38% of the cohort, but this is representative of current state of care. For example, uACR was missing in over 50% of the diabetes population in two large nationally representative datasets.^1,44^ Moreover, our goal was to develop a risk score using real world data from EHR for prediction where uACR is missing in a significant number of patients. More widespread availability of uACR values would enhance the performance of KidneyIntelX, as it was a contributing feature in our model. However, even with this limitation, the performance of KidneyIntelX was more robust than KDIGO strata in those with uACR measured. Second, there was lack of protocolized follow-up resulting in missing data and lack of kidney biopsies, as real-world data from the EHR were used. Missing data can lead to biased machine learning models and the data are prone to ascertainment bias.^45^ However, the median number of eGFR values per patient was 16, and the median time of follow-up was 4.3 years, thereby providing the opportunity to determine whether the kidney outcome was met. Although the primary biobanked cohorts used in the study are broadly representative of the parent hospital populations in terms of age, race/ethnicity and gender distribution and our study is representative of the US DKD population, we cannot rule out an inherent bias since the recruitment was opt-in recruitment and patients who chose to participate in the cohorts from which the study population was selected may be different than those who did not participate in the primary cohorts. Finally, both cohorts are from the Northeast of USA and an independent validation cohort is needed to ensure generalizability. However, only 1/3^rd^ of the participants were white, thus there was adequate representation of racial groups that experience disparities for kidney disease.

In conclusion, a machine learned model combining plasma biomarkers and EHR data significantly improved prediction of progressive decline in kidney function over standard clinical models in patients with T2 DKD from two large academic medical centers.

## Data Availability

Data is available on request and subject to institutional approvals due to patient health information.

## Author Contributions

GNN and FF had full access to all of the data in the study and take responsibility for the integrity of the data and the accuracy of the data analysis.

*Concept and design:* Nadkarni, Fleming, Donovan, Coca

*Acquisition, analysis, or interpretation of data:* Nadkarni, Fleming, Donovan, Coca, Damrauer *Drafting of the manuscript:* Chan, Nadkarni, Coca, Damrauer *Critical revision of the manuscript for important intellectual content:* All authors *Statistical analysis:* Nadkarni, Fleming, Kattan, Coca

*Administrative, technical, or material support:* McCullough, Connolly, Mosoyan *Supervision:* Nadkarni, Fleming, Murphy, Donovan, Coca

## REFERENCES

1. USRDS. USRDS 2018 Annual Data Report: Atlas of Chronic Kidney Disease and End-Stage Renal Disease in the United States, National Institutes of Health, National Institute of Diabetes and Digestive and Kidney Diseases. 2018, 2018.

2. KDIGO. KDIGO 2012 Clinical Practice Guideline for the Evaluation and Management of Chronic Kidney Disease. Kidney Int Suppl.3:1-163.

3. Dunkler D, Gao P, Lee SF, et al. Risk Prediction for Early CKD in Type 2 Diabetes. Clin J Am Soc Nephrol. 2015;10(8):1371–1379.

4. Agrawal V, Ghosh AK, Barnes MA, McCullough PA. Perception of indications for nephrology referral among internal medicine residents: a national online survey. Clin J Am Soc Nephrol. 2009;4(2):323–328.

5. Boulware LE, Troll MU, Jaar BG, Myers DI, Powe NR. Identification and referral of patients with progressive CKD: a national study. Am J Kidney Dis. 2006;48(2):192–204.

6. Hingwala J, Wojciechowski P, Hiebert B, et al. Risk-Based Triage for Nephrology Referrals Using the Kidney Failure Risk Equation. Canadian journal of kidney health and disease. 2017;4:2054358117722782.

7. Kagoma YK, Weir MA, Iansavichus AV, et al. Impact of estimated GFR reporting on patients, clinicians, and health-care systems: a systematic review. Am J Kidney Dis. 2011;57(4):592–601.

8. Sprangers B, Evenepoel P, Vanrenterghem Y. Late referral of patients with chronic kidney disease: no time to waste. Mayo Clin Proc. 2006;81(11):1487–1494.

9. Winkelmayer WC, Liu J, Chertow GM, Tamura MK. Predialysis nephrology care of older patients approaching end-stage renal disease. Arch Intern Med. 2011;171(15):1371–1378.

10. Gillespie BW, Morgenstern H, Hedgeman E, et al. Nephrology care prior to end-stage renal disease and outcomes among new ESRD patients in the USA. Clinical kidney journal. 2015;8(6):772–780.

11. Niewczas MA, Gohda T, Skupien J, et al. Circulating TNF receptors 1 and 2 predict ESRD in type 2 diabetes. J Am Soc Nephrol. 2012;23(3):507–515.

12. Coca SG, Nadkarni GN, Huang Y, et al. Plasma Biomarkers and Kidney Function Decline in Early and Established Diabetic Kidney Disease. J Am Soc Nephrol. 2017;28(9):2786–2793.

13. Looker HC, Colombo M, Hess S, et al. Biomarkers of rapid chronic kidney disease progression in type 2 diabetes. Kidney Int. 2015;88(4):888–896.

14. Damrauer SM, Chaudhary K, Cho JH, et al. Association of the V122I Hereditary Transthyretin Amyloidosis Genetic Variant With Heart Failure Among Individuals of African or Hispanic/Latino Ancestry. JAMA. 2019;322(22):2191–2202.

15. Nadkarni GN, Chauhan K, Verghese DA, et al. Plasma biomarkers are associated with renal outcomes in individuals with APOL1 risk variants. Kidney Int. 2018;93(6):1409–1416.

16. Tayo BO, Teil M, Tong L, et al. Genetic background of patients from a university medical center in Manhattan: implications for personalized medicine. PLoS One. 2011;6(5):e19166.

17. Bajaj A, Ihegword A, Qiu C, et al. Phenome-wide association analysis suggests the APOL1 linked disease spectrum primarily drives kidney-specific pathways. Kidney Int. 2020;97(5):1032–1041.

18. Centers for Disease Control and Prevention (CDC). National Center for Health Statistics (NCHS). National Health and Nutrition Examination Survey Data. Hyattsville, MD: U.S. Department of Health and Human Services, Centers for Disease Control and Prevention, 2018-2019. https://www.cdc.gov/nchs/nhanes/index.htm.

19. Bandelow B, Baldwin DS, Dolberg OT, Andersen HF, Stein DJ. What is the threshold for symptomatic response and remission for major depressive disorder, panic disorder, social anxiety disorder, and generalized anxiety disorder? The Journal of clinical psychiatry. 2006;67(9):1428–1434.

20. Baldwin JA, Johnson RM, Gotz NK, Wayment HA, Elwell K. Perspectives of college students and their primary health care providers on substance abuse screening and intervention. Journal of American college health: J of ACH. 2006;55(2):115–119.

21. Levey AS, Stevens LA, Schmid CH, et al. A new equation to estimate glomerular filtration rate. Ann Intern Med. 2009;150(9):604–612.

22. Leffondre K, Boucquemont J, Tripepi G, Stel VS, Heinze G, Dunkler D. Analysis of risk factors associated with renal function trajectory over time: a comparison of different statistical approaches. Nephrol Dial Transplant. 2015;30(8):1237–1243.

23. Levey AS, Inker LA, Matsushita K, et al. GFR decline as an end point for clinical trials in CKD: a scientific workshop sponsored by the National Kidney Foundation and the US Food and Drug Administration. Am J Kidney Dis. 2014;64(6):821–835.

24. Nelson RG, Grams ME, Ballew SH, et al. Development of Risk Prediction Equations for Incident Chronic Kidney Disease. JAMA. 2019.

25. De Silva AP, Moreno-Betancur M, De Livera AM, Lee KJ, Simpson JA. A comparison of multiple imputation methods for handling missing values in longitudinal data in the presence of a time-varying covariate with a non-linear association with time: a simulation study. BMC Med Res Methodol. 2017;17(1):114.

26. Chicco D. Ten quick tips for machine learning in computational biology. BioData mining. 2017;10:35.

27. Pencina MJ, D’Agostino RB, Sr., D’Agostino RB, Jr., Vasan RS. Evaluating the added predictive ability of a new marker: from area under the ROC curve to reclassification and beyond. Stat Med. 2008;27(2):157–172; discussion 207-112.

28. Pencina MJ, D’Agostino RB, Sr., Steyerberg EW. Extensions of net reclassification improvement calculations to measure usefulness of new biomarkers. Stat Med. 2010;30(1):11–21.

29. Smart NA, Dieberg G, Ladhani M, Titus T. Early referral to specialist nephrology services for preventing the progression to end-stage kidney disease. The Cochrane database of systematic reviews. 2014(6):CD007333.

30. Smart NA, Titus TT. Outcomes of early versus late nephrology referral in chronic kidney disease: a systematic review. Am J Med. 2011;124(11):1073–1080 e1072.

31. Perkovic V, de Zeeuw D, Mahaffey KW, et al. Canagliflozin and renal outcomes in type 2 diabetes: results from the CANVAS Program randomised clinical trials. The lancet. Diabetes & endocrinology. 2018;6(9):691–704.

32. Perkovic V, Jardine MJ, Neal B, et al. Canagliflozin and Renal Outcomes in Type 2 Diabetes and Nephropathy. N Engl J Med. 2019.

33. Kristensen Sl, Rorth R, Jhund PS, et al. Cardiovascular, mortality, and kidney outcomes with GLP-1 receptor agonists in patients with type 2 diabetes: a systematic review and meta-analysis of cardiovascular outcome trials. The lancet. Diabetes & endocrinology. 2019;7(10):776–785.

34. Sarafidis P, Ferro CJ, Morales E, et al. SGLT-2 inhibitors and GLP-1 receptor agonists for nephroprotection and cardioprotection in patients with diabetes mellitus and chronic kidney disease. A consensus statement by the EURECA-m and the DIABESITY working groups of the ERA-EDTA. Nephrol Dial Transplant. 2019;34(2):208–230.

35. Mehrotra R. Advancing American Kidney Health: An Introduction. Clin J Am Soc Nephrol. 2019;14(12):1788.

36. Tummalapalli L, Nadkarni GN, Coca SG. Biomarkers for predicting outcomes in chronic kidney disease. Curr Opin Nephrol Hypertens. 2016;25(6):480–486.

37. Gohda T, Niewczas MA, Ficociello LH, et al. Circulating TNF receptors 1 and 2 predict stage 3 CKD in type 1 diabetes. J Am Soc Nephrol. 2012;23(3):516–524.

38. Krolewski AS, Niewczas MA, Skupien J, et al. Early progressive renal decline precedes the onset of microalbuminuria and its progression to macroalbuminuria. Diabetes Care. 2014;37(1):226–234.

39. Nowak N, Skupien J, Niewczas MA, et al. Increased plasma kidney injury molecule-1 suggests early progressive renal decline in non-proteinuric patients with type 1 diabetes. Kidney Int. 2016;89(2):459–467.

40. Pavkov ME, Nelson RG, Knowler WC, Cheng Y, Krolewski AS, Niewczas MA. Elevation of circulating TNF receptors 1 and 2 increases the risk of end-stage renal disease in American Indians with type 2 diabetes. Kidney Int. 2015;87(4):812–819.

41. Bhatraju PK, Zelnick LR, Shlipak M, Katz R, Kestenbaum B. Association of Soluble TNFR-1 Concentrations with Long-Term Decline in Kidney Function: The Multi-Ethnic Study of Atherosclerosis. J Am Soc Nephrol. 2018;29(11):2713–2721.

42. Tangri N, Grams ME, Levey AS, et al. Multinational Assessment of Accuracy of Equations for Predicting Risk of Kidney Failure: A Meta-analysis. JAMA. 2016;315(2):164–174.

43. Tangri N, Stevens LA, Griffith J, et al. A predictive model for progression of chronic kidney disease to kidney failure. JAMA. 2011;305(15):1553–1559.

44. Tummalapalli SL, Powe NR, Keyhani S. Trends in Quality of Care for Patients with CKD in the United States. Clin J Am Soc Nephrol. 2019;14(8):1142–1150.

45. Gianfrancesco MA, Tamang S, Yazdany J, Schmajuk G. Potential Biases in Machine Learning Algorithms Using Electronic Health Record Data. JAMA Intern Med. 2018;178(11):1544–1547.

